# Increased circulating interleukin concentrations in type 2 diabetes mellitus: A systematic review and meta-analysis

**DOI:** 10.1101/2024.04.13.24305775

**Authors:** Chang Cao, Jing Yuan, Elizabeth R. Gilbert, Mark A. Cline, Fan Lam, King C. Li, Ryan N. Dilger

**Affiliations:** Beckman Institute for Advanced Science and Technology, University of Illinois Urbana-Champaign, Urbana, IL; Department of Animal Sciences, University of Illinois Urbana-Champaign, Urbana, IL; Department of Animal and Poultry Sciences, Virginia Polytechnic Institute and State University, Blacksburg, VA; School of Neuroscience, Virginia Polytechnic Institute and State University, Blacksburg, VA; Department of Bioengineering, University of Illinois Urbana-Champaign, Urbana, IL; Carle-Illinois College of Medicine, University of Illinois Urbana-Champaign, Urbana, IL

## Abstract

**BACKGROUND:** Chronic systemic inflammation links to type 2 diabetes mellitus (T2DM) onset, and the potential role of interleukins in this pathogenic process is increasingly recognized.

**PURPOSE:** To quantitatively evaluate circulating interleukin concentrations data available for T2DM patients.

**DATA SOURCES:** We performed a systematic review using PubMed, Web of Science, and the Cochrane Library.

**STUDY SELECTION:** Original studies reporting data on circulating interleukin concentrations in at least one group of T2DM patients [obese T2DM, overweight T2DM, or lean T2DM] and healthy weight controls (HWCs) were included.

**DATA EXTRACTION AND SYNTHESIS:** Data were extracted from 43 included studies uniquely encompassing 2,646 T2DM patients and 6,605 HWCs by independent investigators. We used a random-effects model to pool data in the Comprehensive Meta-Analysis Version 2 software. Effect sizes were calculated as the standardized mean difference in interleukin concentrations between groups and then transformed into Hedge’s g statistic.

**LIMITATIONS:** Of the ILs included in the analysis (interleukin 2, 4, 6, 8, 10, 12, 17, 18, 22, and 33), 70% were with a high level of between-study heterogeneity and could not be fully accounted for by factors such as BMI, sex, age, and publication year. More studies are warranted to identify more contributing clinical variables.

**CONCLUSIONS:** Meta-analysis outcomes demonstrated higher circulating concentrations of IL-4, IL-6, IL-17, and IL-18 in T2DM patients, strengthening the clinical evidence that T2DM is accompanied by a systemic inflammatory response.

**ARTICLE HIGHLIGHTS:**

- Why did we undertake this study? Possible changes in circulating concentrations for most interleukins (ILs) in type 2 diabetes mellitus (T2DM) patients remains uncertain.
- What is the specific question(s) we wanted to answer? We wanted to quantitatively assess available data on circulating IL concentrations in T2DM patients and healthy weight control (HWC) subjects.
- What did we find? Increased circulating concentrations of IL-4, IL-6, IL-17, and IL-18 were found in T2DM patients compared with HWC.
- What are the implications of our findings? These findings affirm T2DM pathogenesis correlates with systemic inflammation, evidenced by elevated circulating concentrations of both pro- and anti-inflammatory interleukins.

---

The world has witnessed the rising concern of diabetes, a group of chronic endocrine diseases, in recent decades. According to the report from International Diabetes Federation, over 540 million people (aged 20-79 years) around the world have been diagnosed with diabetes. The rate of increase is about 10 million cases per year, resulting in an estimation that there will be approximately 783 million people with diabetes by 2045 (1). In the United States, one out of ten people have diabetes and over 90% of them have type 2 diabetes mellitus (T2DM) (2). Among the common risk factors for T2DM, obesity stands out as a major concern, not only due to its prevalence across all age populations but also the various associated complications, cardiovascular disease, arthritis, and certain types of cancer, to name a few (3). It has long been described that obesity is closely related to a chronic inflammatory subacute state, evidenced by resident immune cells in adipose tissue that secrete inflammatory cytokines to influence systemic concentrations of inflammatory markers (3). Thus, obesity acts as a bridge between T2DM and inflammatory factors, although the relationship between them being reciprocal causation or not remains inconclusive.

Interleukins (ILs) are a group of signaling cytokines (i.e., specialized immune proteins) involved in immune regulation and inflammatory responses. Accumulating evidence has shown that certain interleukins, especially those considered pro-inflammatory, like IL-1β and IL-6, are potentially linked to T2DM (4,5). IL-1β is a key regulator of some critical metabolic processes, affecting insulin secretion and β cells vitality. The balance between macrophage-derived IL-1β concentrations and β-cell-expressed IL-1 receptor antagonist (IL-1Ra) is well-maintained under normal healthy conditions (4). But this balance is gradually broken as obesity and T2DM develop, while IL-1β concentration becomes chronically elevated (4). In this context, patients with obesity or T2DM are supposed to have increased IL-1β compared with healthy controls. However, the difference is not statistically significant based on a previous meta-analysis (4). Compared to IL-1β, IL-6 has been more widely studied, suggesting a connection between elevated IL-6 concentrations and higher T2DM incidence (6). Previous systematic and meta-analyses on IL-6 have thus far predominantly focused on populations from North America and Europe, while risks associated with inflammatory conditions remain a global concern (5–7). Besides these two ILs, other pro-inflammatory ILs, such as IL-8 and IL-17, and anti-inflammatory ILs, like IL-4 and IL-10, have not been thoroughly studied, leaving our understanding of their relationship with T2DM risk remaining insufficient.

Body mass index is a well-accepted measure drawing people’s attention to take caution and triggering their action when approaching the cutoffs for overweight and obesity (25 and 30 kg/m^2^, respectively, according to the World Health Organization) (8). Higher BMI correlates with increased T2DM risk, particularly with central obesity. Abdominal visceral fat, a major source of inflammatory cytokines, exacerbates this risk (3). While many studies use BMI generically, few stratify T2DM population by BMI to explore aberration of inflammatory cytokine concentrations. Despite the nonlinear BMI-T2DM risk relationship, even those with healthy weight can develop T2DM. Weight loss remains effective in risk reduction (9). Thus, BMI, as a clinical screening tool, can promote a better understanding of various T2DM stages and T2DM-related research scenarios.

Our current study entailed a systematic review and meta-analysis comparing circulating interleukin concentrations in T2DM patients and healthy weight controls (HWCs), with BMI as a major moderator. Subgroups based on BMI were assessed to evaluate its impact on interleukin concentrations. We considered and investigated the effects of sex, age, and publication year to account for potential confounding variables that could complicate interpretation of findings.

## METHODS

Our systematic review and meta-analysis protocol was registered with International prospective Register of Systematic Reviews (PROSPERO, registration no. CRD42024522443) and conducted following the Preferred Reporting Items for Systematic reviews and Meta-Analysis (PRISMA) guidelines (**Supplemental Table S3**)(10).

### Search strategy and exclusion criteria

Two researchers (C.C. and J.Y.) independently conducted a systematic review of English-language articles, using PubMed, Web of Science, and the Cochrane Library. The last search was performed in March 2023. The search query used was: [interleukins OR their abbreviation (e.g., interleukin 1 OR IL-1)] AND [type 2 diabetes] AND [obesity OR obese OR overweight]. We searched for 23 different ILs (i.e., interleukin 1, 2, 4, 5, 6, 7, 8, 9, 10, 12, 13, 15, 17, 18, 21, 22, 23, 27, 28, 31, 33, 35, and 37). There were no restrictions on publication year. Initially, the search yielded 2,548 records from PubMed, 9,658 from Web of Science, and 144 from the Cochrane Library. After titles and abstracts scanning, 197 relevant publications were chosen for full-text review. Inclusion criteria sought original studies reporting circulating interleukin concentrations in at least one group of T2DM patients [obese T2DM (OBD), overweight T2DM (OWD), OR lean T2DM (LD)] and HWC. Out of 197 articles, 154 were excluded for various reasons, including lacking necessary data (such as no BMI or IL concentrations data), reporting data for only one group without a comparative HWC, measuring interleukin concentrations in peripheral blood cells. Additionally, studies involving patients with other diseases or healthy subjects with a family history of T2DM and in vitro studies were excluded, as were studies where patients were not diagnosed with T2DM at the time of blood sampling, and studies that assessed interleukins concentrations being less than three. Consequently, 43 publications were included in the present meta-analysis (11–53) (PRISMA flowchart, **Figure 1**), covering 10 different ILs (interleukin 2, 4, 6, 8, 10, 12, 17, 18, 22, and 33).

**Figure 1.**
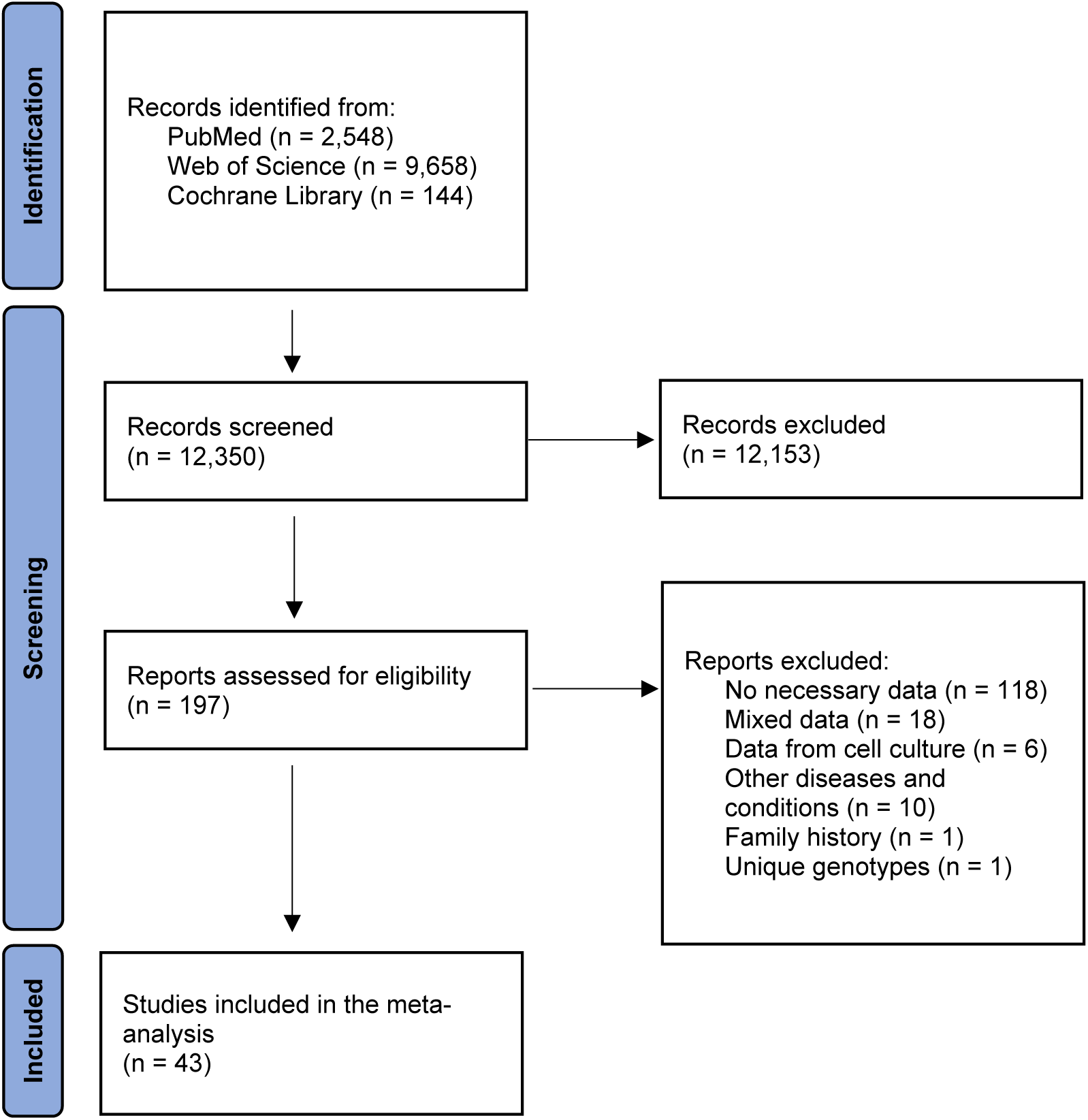
PRISMA flowchart of the systematic literature search. A systematic search of PubMed, Web of Science, and the Cochrane Library ultimately identified 43 articles to be included in the meta-analysis.

### Data extraction

We gathered information concerning various aspects, including sample size, the average or median interleukin concentrations, standard deviation (SD) and *P*-value as our primary focus. Additionally, we collected data on patient demographics including mean age, BMI, and sex distribution, as well as diabetic diagnosis criteria and duration, blood fraction analyzed (serum or plasma), study location (country), patient medications and any existing comorbidity, and measurement techniques. To ensure the accuracy of the data extraction, one investigator (C.C.) compiled the information, which was then cross-verified by another investigator (J.Y.), and any discrepancies were resolved through consensus. Included studies, along with demographic and clinical characteristics, are summarized in **Supplemental Table S1**. It is worth noting that some studies examined the influence of medications, exercise, or other diseases, but baseline interleukin concentrations were measured at enrollment, or patients using other medications assumed to have no effect on interleukin concentrations, in which case the data was included in the meta-analysis.

### Statistical analysis

We conducted all statistical analyses using Comprehensive Meta-Analysis Version 2 software (Biostat, Englewood, NJ, USA) (54). As BMI is a major moderating factor we intended to explore, the following analyses were conducted both on overall and individual BMI subgroups when feasible. Effective sizes (ESs) were computed primarily using sample size, mean values, and SD. In cases of missing mean and SD data, we used sample size and *P*-values to compute ESs. The ESs were calculated as the standardized mean difference in interleukin concentrations between groups and then transformed into Hedge’s g, which offers an unbiased ES adjusted for sample size. We employed 95% CI to evaluate pooled ES statistical significance. For our meta-analysis, we opted for random-effects models, anticipating differences in the true ES due to within-study and between-study variables (54). To ensure the robustness of our findings, we ran the sensitivity analysis, which systematically excluded one study at a time to check if the results of the meta-analysis were stable (data not shown).

We assessed the statistical difference of between-study heterogeneity using the Cochran Q test, with a significance level set at *P*-value < 0.1. The I^2^ index was used to measure the inconsistency across studies and gauge the extent of heterogeneity, with I^2^ values of 0.25, 0.50, and 0.75 serving as thresholds for low, moderate, and high levels of heterogeneity, respectively. We also performed random-effects meta-regressions of ES, employing unrestricted maximum-likelihood models to explore whether covariates such as mean age, sex distribution, and the publication year of each study acted as moderators influencing the ES. To visually assess publication bias, we used funnel plots, plotting the ES against the precision (i.e., inverse of the standard error) for each study (data not shown). The Egger’s test was employed to statistically test for publication bias by assessing funnel plot asymmetry. Additionally, we utilized the Classic Fail-Safe N analysis to estimate the number of missing (i.e., unpublished) studies required to raise the observed *P*-value to greater than 0.05, thereby suggesting potential publication bias (data not shown).

In all cases, statistical significance was accepted as *P*-value < 0.05 unless otherwise specified, with 0.05 < *P*-value < 0.1 reported as a trend.

## RESULTS

### Characteristics of the identified studies

We conducted a systematic search that yielded 2,548 records from PubMed, 9,658 from Web of Science, and 144 from the Cochrane Library. Following a review of the titles and abstracts, we identified 197 articles that were related to our meta-analysis and subjected them to a full-text examination. Ultimately, we included a total of 43 studies (illustrated in **Figure 1**), encompassing 2,646 patients with T2DM and 6,605 HWCs. Most of these studies focused on obese diabetic patients (N = 53%), while overweight and lean diabetic patients made up 35% and 21% of the entire patient cohort, respectively. Some studies had multiple BMI subgroups, causing the overall percentage to exceed 100% (**Table 1**). The sample sizes for T2DM patients ranged from 7 to 476, and 8 to 2,756 for HWCs. The mean age of participants ranged from 32.4 to 71.5 years for T2DM patients and from 26.0 to 66.0 years for HWCs (**Supplemental Table S1**).

**Table 1.** Summary of comparative outcomes for circulating interleukin measurements

| Interleukin | Group | No. of Studies | No. with T2DM/HWC | Main effect |  |  | Heterogeneity |  |  | Publication Bias |  |  |
| --- | --- | --- | --- | --- | --- | --- | --- | --- | --- | --- | --- | --- |
|  |  |  |  | Hedges g (95% CI) | Z Score | P-value | Q Statistic | df | P-value | I<sup>2</sup> Statistic | Egger Intercept | P-value |
| IL-6 | Overall | 28 | 1771/3345 | 1.317 ( 1.045 to 1.588 ) | 9.507 | **<0.001** | 239.183 | 27 | <0.001 | 88.712 | 1.06205 | 0.243 |
|  | *OBD* | 16 | 806/381 | 1.736 ( 1.200 to 2.273 ) | 6.341 | **<0.001** | 151.960 | 15 | <0.001 | 90.129 | 6.69923 | <0.001 |
|  | *OWD* | 8 | 705/2902 | 1.257 ( 0.791 to 1.723 ) | 5.284 | **<0.001** | 86.731 | 7 | <0.001 | 91.929 | 0.43853 | 0.836 |
|  | *LD* | 7 | 260/240 | 0.575 ( 0.386 to 0.764 ) | 5.974 | **<0.001** | 5.431 | 6 | 0.490 | 0.000 | -0.16876 | 0.918 |
| IL-18 | Overall | 9 | 333/262 | 1.660 ( 0.910 to 2.409 ) | 4.341 | **<0.001** | 112.462 | 8 | <0.001 | 92.886 | 5.88082 | 0.077 |
|  | *OBD* | 6 | 162/110 | 2.237 ( 1.033 to 3.440 ) | 3.642 | **<0.001** | 66.509 | 5 | <0.001 | 92.482 | 10.30397 | 0.030 |
|  | *OWD* | 3 | 171/152 | 0.719 ( -0.213 to 1.650 ) | 1.512 | 0.130 | 30.847 | 2 | <0.001 | 93.516 | -25.54789 | 0.394 |
| IL-10 | Overall | 7 | 225/233 | 0.310 ( -1.509 to 2.128 ) | 0.334 | 0.739 | 340.784 | 6 | <0.001 | 98.239 | -10.52366 | 0.519 |
|  | *OBD* | 5 | 185/186 | 0.407 ( -2.261 to 3.076 ) | 0.299 | 0.765 | 334.709 | 4 | <0.001 | 98.805 | -17.27322 | 0.547 |
| IL-17 | Overall | 6 | 244/184 | 2.538 ( 0.692 to 4.383 ) | 2.695 | **0.007** | 244.620 | 5 | <0.001 | 97.956 | 12.40433 | 0.092 |
|  | *LD* | 3 | 160/92 | 2.954 ( 0.369 to 5.540 ) | 2.239 | **0.025** | 90.819 | 2 | <0.001 | 97.798 | 15.41625 | 0.180 |
| IL-2 | Overall | 4 | 142/146 | 0.316 ( -0.081 to 0.713 ) | 1.562 | 0.118 | 7.447 | 3 | 0.059 | 59.717 | 4.70571 | 0.061 |
| IL-4 | Overall | 4 | 196/191 | 0.675 ( 0.072 to 1.278 ) | 2.193 | **0.028** | 22.446 | 3 | <0.001 | 86.634 | -2.33836 | 0.755 |
|  | *OWD* | 3 | 168/179 | 0.754 ( 0.014 to 1.494 ) | 1.996 | **0.046** | 21.291 | 2 | <0.001 | 90.607 | -1.74124 | 0.934 |
| IL-8 | Overall | 4 | 130/66 | 0.235 ( -0.241 to 0.711 ) | 0.968 | 0.333 | 6.880 | 3 | 0.076 | 56.398 | -0.73431 | 0.906 |
|  | *OBD* | 3 | 75/47 | 0.487 ( 0.119 to 0.856 ) | 2.590 | **0.010** | 0.520 | 2 | 0.771 | 0.000 | 0.34339 | 0.925 |
| IL-12 | Overall | 4 | 205/218 | 1.198 ( -0.050 to 2.447 ) | 1.881 | 0.060 | 93.810 | 3 | <0.001 | 96.802 | 6.87957 | 0.691 |
| IL-22 | Overall | 3 | 395/2806 | 0.844 ( -0.321 to 2.009 ) | 1.421 | 0.155 | 56.512 | 2 | <0.001 | 96.461 | 7.29718 | 0.043 |
| IL-33 | Overall | 3 | 118/125 | -0.172 ( -0.659 to 0.315 ) | -0.692 | 0.489 | 6.317 | 2 | 0.042 | 68.342 | 6.02978 | 0.104 |

Abbreviation: No., number; T2DM, type 2 diabetes mellitus; HWC, healthy weight control; IL, interleukin; OBD, obese type 2 diabetes mellitus; OWD, overweight type 2 diabetes mellitus; LD, lean type 2 diabetes mellitus; CI, confidence interval; df, degree of freedom; Q, Cochran's Q test; Z (test of null hypothesis); *P* (statistical significance); I<sup>2</sup> (heterogeneity level).

### Meta-analysis of interleukin concentrations

We examined 10 different interleukins between patients with T2DM and HWCs. Our random-effects meta-analysis revealed that individuals with T2DM exhibited significantly higher circulating interleukin concentrations when compared with HWCs (**Table 1**; **Figures 2&3**), including IL-4 (Hedges g, 0.675; 95% CI, 0.072-1.278; *P* = 0.028), IL-6 (Hedges g, 1.317; 95% CI, 1.045-1.588; *P* < 0.001), IL-17 (Hedges g, 2.538; 95% CI, 0.692-4.383; *P* = 0.007), and IL-18 (Hedges g, 1.660; 95% CI, 0.910-2.409; *P* < 0.001). Moreover, T2DM patients, rather than HWCs, showed a tendency towards increased concentrations of IL-12 (Hedges g, 1.198; 95% CI, -0.050 to 2.447; *P* = 0.060; **Table 1**). However, concentrations of IL-2 (Hedges g, 0.316; 95% CI, -0.081 to 0.713; *P* = 0.118), IL-8 (Hedges g, 0.235; 95% CI, -0.241 to 0.711; *P* = 0.333), IL-10 (Hedges g, 0.310; 95% CI, -1.509 to 2.128; *P* = 0.739), IL-22 (Hedges g, 0.844; 95% CI, -0.321 to 2.009; *P* = 0.155), and IL-33 (Hedges g, -0.172; 95% CI, -0.659 to 0.315; *P* = 0.489) did not yield significant ES estimates (**Table 1**).

**Figure 2.**
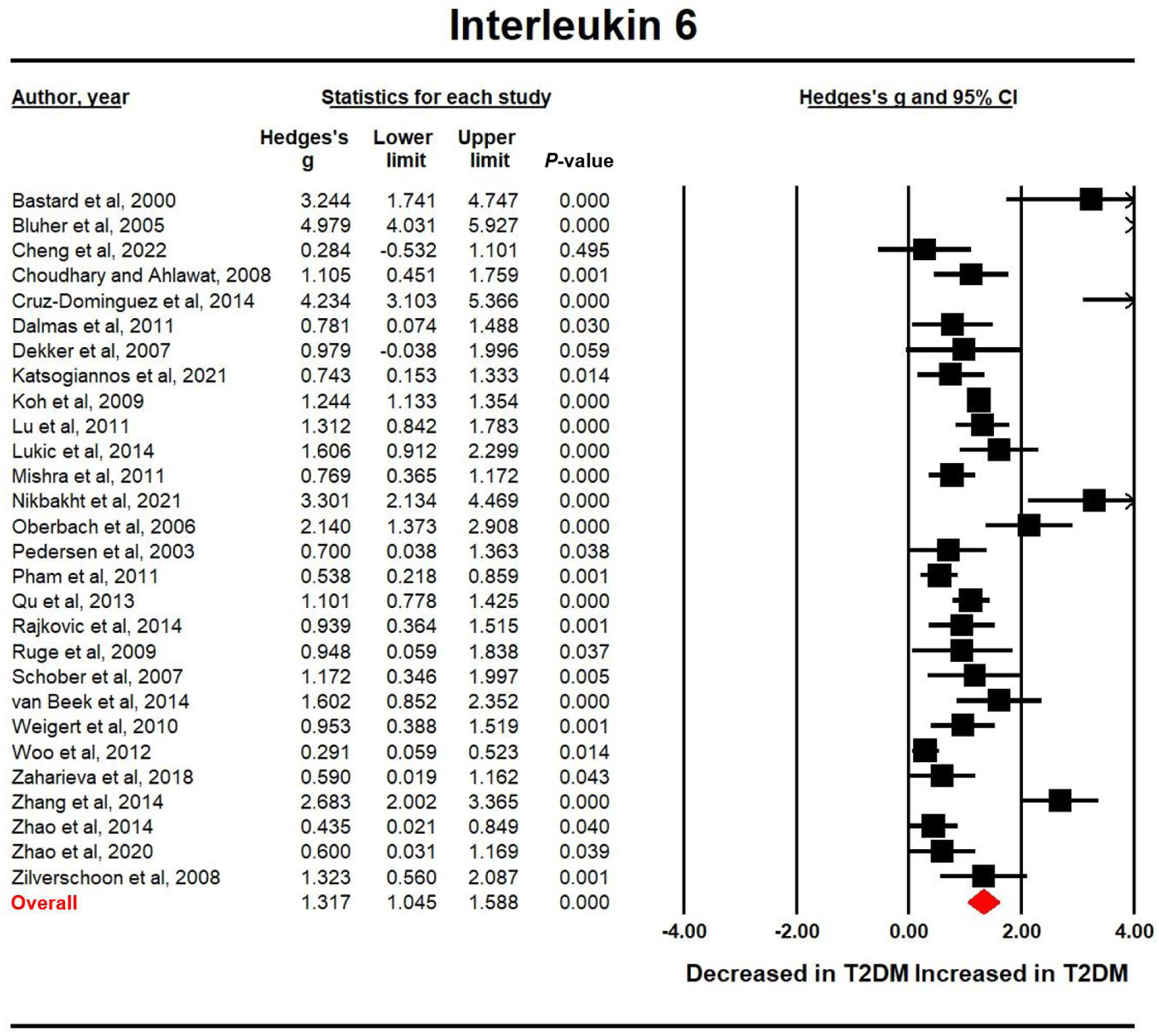
Studies of circulating interleukin-6 concentrations. Forest plot displays random-effects meta-analysis outcomes of the association between interleukin-6 concentrations and type 2 diabetes mellitus (T2DM). The sizes of the squares are proportional to study weights.

**Figure 3.**
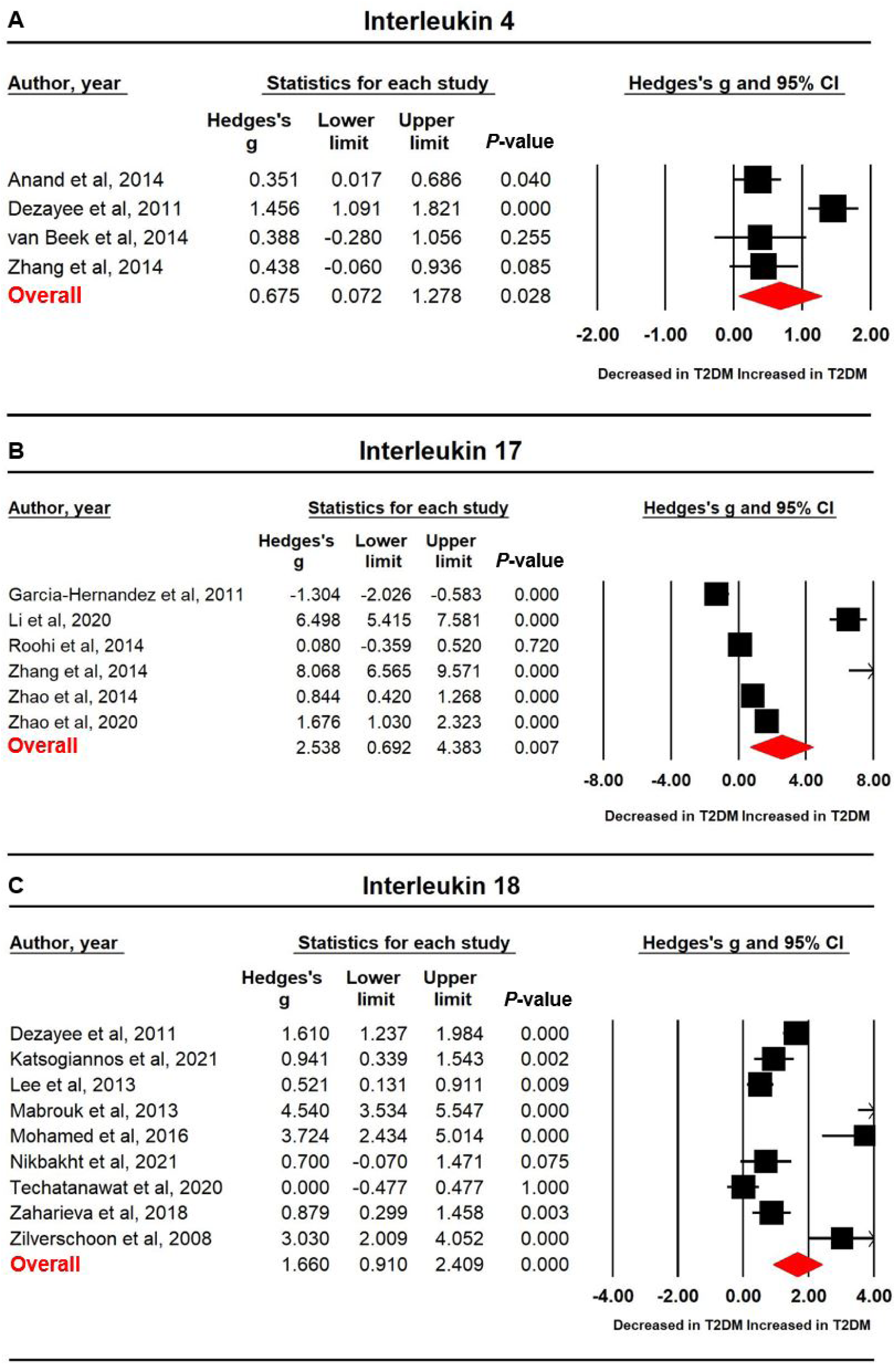
Studies of circulating concentrations of interleukin-4, interleukin-17, and interleukin-18. Forest plot displays random-effects meta-analysis outcomes of the association between interleukin-4, interleukin-17, and interleukin-18 concentrations and type 2 diabetes mellitus (T2DM). The sizes of the squares are proportional to study weights.

Regarding BMI subgroups (**Table 1**), it was unsurprising to observe that all three subgroups of T2DM patients displayed elevated IL-6 concentrations compared with healthy controls, with OBD patients having an even higher ES (Hedges g, 1.736; 95% CI, 1.200-2.273; *P* < 0.001) than moderate level in OWD (Hedges g, 1.257; 95% CI, 0.791-1.723; *P* < 0.001) and low level in LD (Hedges g, 0.575; 95% CI, 0.386-0.764; *P* < 0.001). Similar to the overall results, higher interleukin concentrations in different BMI subgroups of T2DM patients were observed compared with HWCs in IL-4 (OWD, Hedges g, 0.754; 95% CI, 0.014-1.494; *P* = 0.046), IL-17 (LD, Hedges g, 2.954; 95% CI, 0.369-5.540; *P* = 0.025), and IL-18 (OBD, but not OWD, Hedges g, 2.237; 95% CI, 1.033-3.440; *P* < 0.001). For IL-8, OBD patients were found to have a higher concentration than HWCs (Hedges g, 0.487; 95% CI, 0.119-0.856; *P* = 0.010), although this effect did not apply to all patients with T2DM.

### Investigation of heterogeneity

High levels of heterogeneity were observed for 7 of the 10 interleukins, while the remaining three (IL-2, IL-8, and IL-33) displayed moderate levels of heterogeneity (**Table 1**). Thus, we performed meta-regression analyses to account for potential factors that could explain the heterogeneity observed in current meta-analysis, as shown in **Supplemental Table S2**. The moderators we considered included sex, age, and publication year. Due to limited data and study availability for most interleukins, factors such as country where the study was conducted, mean duration since T2DM diagnosis, medication status, and measurement types were not included in the meta-regression analyses.

For IL-6, the impact of heterogeneity decreased to 0% (Q_6_ = 5.431; *P* = 0.490; I^2^ < 0.001) in LD patients compared with the impact in overall T2DM patients (Q_27_ = 239.183; *P* < 0.001; I^2^ = 88.712) (**Table 1**). Interestingly, none of the moderators (sex, age, or publication year) were found to influence the results of the meta-analysis for IL-6 (**Supplemental Table S2**). For IL-8, a similar reduction of the impact of heterogeneity was found in OBD patients (Q_2_ = 0.520; *P* = 0.771; I^2^ < 0.001) versus overall T2DM patients (Q_3_ = 6.880; *P* = 0.076; I^2^ = 56.398) (**Table 1**). We found a significant association between age and ES (coefficient [95% CI], -0.05854 [-0.11458 to -0.00250]; *P* = 0.041), whereas no associations were found between sex or publication year and ES (**Supplemental Table S2**).

For overall T2DM patients, meta-regression analyses revealed that sex was linked to ES in studies measuring IL-12, IL-17, and IL-22 concentrations, whereas age was a major moderator for studies measuring IL-10, IL-17, IL-18, and IL-22 concentrations. Publication year was found to affect ES for studies measuring IL-4, IL-12, and IL-33 concentrations. In various BMI subgroups, all three moderators were found to be linked with ES in studies evaluating IL-4 concentrations in OWD patients and IL-10 concentrations in OBD patients. For studies reporting IL-18 concentrations, age and publication year had an impact on ES in both OBD and OWD patients. Detailed analysis results are available in **Supplemental Table S2**.

### Publication bias

We evaluated publication bias both through quantitative methods involving linear regression analysis and by visually inspecting funnel plots for each interleukin and BMI group. The results indicated the absence of significant publication bias for most interleukins, with the exception of IL-22 (*P* = 0.043, **Table 1**). Likewise, for most of the BMI subgroups, we did not detect any notable publication bias, except for IL-6 and IL-18 in OBD patients (*P* < 0.001 and *P* = 0.030, respectively, **Table 1**).

## DISCUSSION

We carried out a comprehensive systematic review and meta-analysis in comparing circulating interleukins in T2DM patients and HWCs, and considered potential influencing factors like BMI, age, and sex. Our study synthesized data from 43 studies, encompassing 2,646 patients with T2DM and 6,605 HWCs. We investigated most of the identified interleukins and observed significant increases in circulating concentrations of IL-4, IL-6, IL-17, and IL-18 in T2DM patients compared with HWCs. Notably, the ES for IL-17 was the largest, while the ES for IL-4, IL-6, and IL-18 ranged from small to moderate. Sensitivity analyses suggested that the associations observed for IL-6 and IL-18 concentrations and their link to T2DM remained robust and were not significantly impacted by any single study. However, the significant association found in IL-17, IL-4, and several other interleukins listed in Table 1 could be affected by a single study, probably due to the large ES in one study (50) for IL-17, and the limited number of studies for IL-4 and other interleukins. Despite some inconsistency in clinical data across individual interleukins and across studies, the results from current meta-analysis offer strong clinical evidence of an elevated interleukin profile in T2DM patients.

Over the past two decades, the chronic systematic inflammatory state has emerged as a major risk factor for obesity and T2DM (55). The activation and elevation of inflammatory cytokines and adipokines, especially interleukins as the focus of the current study, are thought to be part of the pathogenesis of T2DM (56). IL-1, a pro-inflammatory cytokine, is known to disrupt insulin signaling and contribute to disease progression. However, due to a limited number of available studies, we were unable to conduct meta-analyses on IL-1 and IL-1Ra concentrations in T2DM, although their concentrations consistently appeared higher in T2DM patients compared with HWCs (data not shown, < 3 studies available). Therefore, further research is necessary to establish more robust evidence regarding circulating IL-1 and its receptor concentrations. Previous meta-analyses on IL-6 concentrations in T2DM indicated a significant association between elevated IL-6 concentrations and a higher risk of developing T2DM (5–7). Our findings align with these studies, suggesting that T2DM patients exhibited higher circulating concentrations of IL-6 compared with HWCs. In comparison to earlier investigations, we included about twice as many qualified studies, enhancing the statistical power of our analysis to provide stronger evidence of the IL-6 profile. This increased strength may be attributed to new evidence, more tested populations from regions like Asia and Africa, and our method of converting the mean difference in IL-6 concentrations between T2DM patients and HWCs into an unbiased ES. The current meta-analysis on IL-18 also provided consistent, but more robust, results than a previous study (7), underscoring that T2DM patients had higher concentrations of circulating IL-18 than HWCs. IL-18 belongs to the IL-1 family, also serving as a pro-inflammatory factor and a metabolism mediator in the development of obesity and T2DM (57). Prior meta-analyses on IL-17 concentrations in T2DM yielded inconsistent results (58,59), but our current analysis supported the more recent finding that IL-17 concentrations significantly increased in T2DM patients with no apparent change in HWCs. Like IL-18, IL-17 is a versatile inflammatory marker that mediates adipocyte maturation and function, and regulates the expressions of other pro-inflammatory cytokines, such as IL-6, which might propagate obesity and initiate T2DM development (57).

The two well-known anti-inflammatory interleukins, IL-4 and IL-10, presented divergent results in our meta-analyses. IL-4 showed higher circulating concentrations in T2DM patients, while IL-10 exhibited no difference. Previous meta-analyses reported both decreased and increased serum concentrations of IL-10 (58,59), and our findings, along with prior studies, do not provide a consensus. These discrepancies may stem from variations in study designs and analytical approaches, warranting further investigations to elucidate the relationship between IL-10 and T2DM. Our meta-analysis on IL-4 may be the first undertaken to explore changes in its circulating concentrations in T2DM patients. While only 4 qualified studies were included, the meta-analysis revealed that T2DM patients had elevated IL-4 concentrations compared with HWCs. Besides its capability to counteract inflammation, IL-4 is known to prevent adipogenesis, promote lipolysis, and improve insulin sensitivity (57). The increased circulating concentrations of IL-4 in T2DM patients suggest a mechanism combating the chronic inflammatory state during the development of obesity and T2DM. The observed trend of increasing concentrations of circulating IL-12 in our study potentially supports the role of IL-12 in both early and systemic inflammatory responses in obesity and T2DM (57). Nonetheless, more studies are needed to validate this finding.

Since the 1990s, BMI has been a widely recognized method to categorize surplus weight in individuals (8). A recent study has even emphasized the necessity to lower the cutoff lines for specific populations in areas such as South Asia and Africa, as the existing cutoffs were established based on White populations in industrialized countries (8). To keep it consistent, however, we still used the traditional cutoffs in the current study but acknowledge the critical role of BMI in overall interpretation of results. As such, we divided the T2DM patients into three subgroups, OBD, OWD, and LD, and subsequently conducted meta-analyses for each IL within each BMI subgroup. Not surprisingly, each subgroup showed higher concentrations of IL-6 in T2DM patients compared with HWCs, likely indicating a pivotal role for IL-6 in T2DM development. For other ILs, due to the limited number of studies included, increased circulating concentrations were only found in OBD for IL-18, LD for IL-17, and OWD for IL-4. Thus, we cannot conclude that these increases are exclusive to these subgroups for the aforementioned ILs. However, at least for IL-18, BMI appears to be a significant impact factor, as OWD individuals did not exhibit higher concentrations of this pro-inflammatory interleukin. Interestingly, when we examined IL-8 concentrations in OBD patients, they were significantly higher than those in HWCs. The pleiotropic roles of IL-8 in metabolic inflammation and its association with T2DM were extensively described in a prior study (60). Although not included in our meta-analysis due to its control group being obese individuals, this study paved the way for us to identify that IL-8 could be a novel biomarker for early diagnosis and prevention of T2DM at least in obese population.

Interestingly, the heterogeneity for IL-6 in LD patients and IL-8 in OBD patients dropped to zero from a moderate-to-high level when all T2DM patients were compared with HWCs. Furthermore, higher ESs were consistently found in the OBD subgroup compared with HWCs in our meta-analysis. These findings not only demonstrated that BMI, as a potential confounder, could partially explain the high-level between-study heterogeneity, but underscored the robustness of our meta-analysis with BMI-based subgroup stratification. High levels of heterogeneity were found in those interleukins (IL-4, IL-17, and IL-18) significantly associated with T2DM. Due to the limited availability of studies, we were unable to conduct meta-regression analyses on possible confounders such as country, diabetic duration, and measurement types. However, three continuous variables, sex distribution, mean age of T2DM patients, and paper publication year, stood out as moderators impacting the results of our meta-analysis. Nevertheless, the possibility that their impacts were secondary to those clinical variables like diabetic state and medication duration cannot be ruled out. Thus, future studies investigating the relationship between circulating cytokines and T2DM should include information about these clinical variables.

The current meta-analysis, while providing compelling evidence of strong positive associations between circulating IL concentrations and incidence of T2DM, has some limitations. First, the lack of critical clinical variables hindered a more in-depth exploration of the significant heterogeneity between studies. Second, despite a comprehensive screening of most identified interleukins (23 in total), we could not conduct meta-analysis or identify statistically significant changes in their circulating concentrations in T2DM patients compared with HWCs due to the limited number of studies or participants, as observed with IL-1, IL-2, and IL-12. Third, BMI does not take into account factors like muscle mass, fat distribution, or individual variations in body composition. Therefore, although BMI has historically served as a practical clinical measurement, a more comprehensive assessment of possible moderators is essential when estimating the risk of T2DM development. Fourth, because the meta-analysis provided pooled results from cross-sectional studies, it remained unknown whether the increase in circulating IL concentrations was causally related (as a cause or consequence) to T2DM development. Many researchers have attempted to reveal this relationship by investigating IL concentrations in various types of cells or tissues from which ILs originate (58,59). But there is limited evidence available from a longitudinal perspective, with one exemplary dataset that estimated serum IL-1Ra concentrations longitudinally from years before diagnosis through the conclusion of follow-up (61), suggesting preventive and therapeutic value of investigating IL-blocking agents. These limitations underscore the necessity for ongoing research into the variations in circulating interleukins in T2DM.

## CONCLUSIONS

In summary, this meta-analysis demonstrated elevated circulating interleukins IL-4, IL-6, IL-17, and IL-18 concentrations in T2DM individuals compared with HWCs. Results pertaining to those pro-inflammatory interleukins reinforce the strong association between systemic inflammation and development of obesity and T2DM, while the discovery of increased anti-inflammatory IL-4 concentrations in T2DM patients suggests the presence of a self-regulating mechanism within the body to counteract or balance the inflammatory response. Among the factors affecting the circulating IL concentrations and T2DM development, body weight is an important one and can be easily calculated as BMI index, which has historically provided a practical tool to monitor health status on a daily basis. With the prospect of future studies focusing on interleukin-related inflammation, potentially involving international collaboration, blood interleukins could evolve into crucial biomarkers and therapeutic targets benefiting the prevention and treatment of T2DM.

## Data Availability

All data produced in the present study are available upon reasonable request to the authors

## ACKNOWLEDGEMENTS

Funding. C.C. is supported by the Postdoctoral Fellows Program via Beckman Institute for Advanced Science and Technology at University of Illinois Urbana-Champaign.

The funder had no role in considering the study design or in the collection, analysis, or interpretation of data, writing of the report, or decision to submit the manuscript for publication.

## Author Contributions

C.C., E.R.G., and M.A.C. conceptualized the study. C.C. and R.N.D. were involved in the study design. C.C. and J.Y. conducted the systematic search, study selection, and data extraction. C.C. performed the formal statistical analyses and interpretation of the results. J.Y. validated the data analysis and result interpretation. C.C. wrote the first draft of the manuscript, and all authors (C.C., J.Y., E.R.G., M.A.C., F.L., K.C.L., and R.N.D.) edited, reviewed, and approved the final version of the manuscript. C.C. is the guarantor of this work and, as such, had full access to all the data in the study and takes responsibility for the integrity of the data and the accuracy of the data analysis.

