## Supplemental Tables for "Increased circulating interleukin concentrations in type 2 diabetes mellitus: A systematic review and meta-analysis"

**Table of Contents**

**Supplemental Tables**

Supplemental Table S1. Characteristics of included studies evaluating interleukin concentrations

Supplemental Table S2. Meta-regression analyses

Supplemental Table S3. PRISMA 2020 Checklist

Supplemental Table S1. Characteristics of included studies evaluating interleukin concentrations

| Author/year | Interleukin | No. (T2DM/ HWC) | Sex (% Male) (T2DM/ HWC) | Age (years) (T2DM/ HWC) | BMI (T2DM/ HWC) | Sample Origin | Measurement | Country | Reported Medication | Reported Comorbidity | Diabetes diagnosis criteria | Diabetes duration (years) | Fasting |
| --- | --- | --- | --- | --- | --- | --- | --- | --- | --- | --- | --- | --- | --- |
| Anand/ 2014 | IL-2, IL-4, IL-12, IL-33 | OWD 61/ HWC 79 | OWD 62/ HWC 51 | OWD 54.7/ HWC 46.2 | OWD 25.2/ HWC 22.9 | Serum | Bio-Plex multiplex cytokine bead assay (IL-2, IL-4, IL-12), ELISA (IL-33) | India | Y | N | ADA | NA | Y |
| Barchetta/ 2017 | IL-8 | OBD 35/ OWD 36/ HWC 21 | OBD 74/ OWD 58/ HWC 67 | OBD 51.9/ OWD 50.4/ HWC 52.3 | OBD 34.2/ OWD 26.6/ HWC 24.4 | Serum | Bio-Plex multiplex cytokine bead assay | Italy | N | N | ADA | OBD 6.2/ OWD 7.6 | Y |
| Bastard/ 2000 | IL-6 | OBD 7/ HWC 8 | OBD 0/ HWC 0 | OBD 58/ HWC 42 | OBD 36.6/ HWC 20.6 | Serum | ELISA | France | NA | N | ADA | NA | Y |
| Bluher/ 2005 | IL-6, IL-10 | OBD 26/ HWC 45 | OBD 54/ HWC 49 | NA | OBD 31.6/ HWC 24.2 | Serum | ELISA | Germany | Y | N | ADA | NA | Y |
| Cakici/ 2021 | IL-33 | OBD 36/ HWC 21 | NA | NA | OBD 33.65/ HWC 23.44 | Serum | ELISA | Turkey | NA | N | ADA | NA | Y |
| Cheng/ 2021 | IL-22 | OWD 275/ HWC 2756 | OWD 40/ HWC 32 | OWD 59.7/ HWC 52.2 | OWD 25.2/ HWC 24.0 | Plasma | ELISA | China | N | N | ADA | NA | Y |
| Cheng/ 2022 | IL-6, IL-10 | LD 8/ HWC 17 | LD 25/ HWC 29 | LD 70.0/ HWC 66.0 | LD 23.0/ HWC 20.7 | Serum | ELISA | China | N | N | WHO | NA | Y |
| Choudhary and Ahlawat/ 2008 | IL-6 | LD 20/ HWC 20 | LD 50/ HWC 50 | LD 54.25/ HWC 55.5 | LD 23.36/ HWC 23.74 | Serum | ELISA | India | N | N | ADA | 9.45 | Y |
| Cruz-Dominguez/ 2014 | IL-6 | OBD 9/ HWC 36 | OBD 11/ HWC 50 | OBD 41.75/ HWC 38.00 | OBD 45.9/ HWC 24.0 | Plasma | ELISA | Mexico | N | N | ADA | NA | Y |
| Dalmas/ 2011 | IL-6 | OBD 18/ HWC 14 | OBD 0/ HWC 0 | OBD 47.9/ HWC 38.6 | OBD 52.8/ HWC 21.5 | Serum | ELISA | France | Y | Y | WHO | NA | Y |
| Dekker/ 2007 | IL-6 | OWD 8/ HWC 8 | OWD 100/ HWC 100 | OWD 51.0/ HWC 47.5 | OWD 29.9/ HWC 24.5 | Serum | ELISA | Canada | Y | N | WHO | NA | Y |
| Dezayee/ 2011 | IL-4, IL-12, IL-18 | OWD 75/ HWC 70 | OWD 33/ HWC 54 | OWD 50.0/ HWC 45.3 | OWD 27.78/ HWC 23.63 | Serum | ELISA | Iraq | Y | N | NA | 8.21 | Y |
| García-Hernández/ 2011 | IL-17 | OBD 14/ HWC 22 | OBD 29/ HWC 32 | OBD 47.0/ HWC 31.0 | OBD 31.9/ HWC 21.4 | Serum | ELISA | Mexico | Y | N | ADA | NA | Y |
| Hang/ 2014 | IL-8, IL-12 | LD 19/ HWC 19 | LD 47/ HWC 63 | LD 64.1/ HWC 62.9 | LD 23.0/ HWC 21.9 | Plasma | Milliplex human cytokine/chemokine panel | China | NA | N | NA | 9.0 | NA |
| Katsogiannos/ 2021 | IL-2, IL-6, IL-18, IL-33 | OBD 21/ HWC 25 | OBD 14/ HWC 44 | OBD 49/ HWC 39 | OBD 38.3/ HWC 24.4 | Plasma | Magnetic bead-based Luminex assay | Sweden | Y | NA | NA | 5 | Y |
| Koh/ 2009 | IL-6 | OWD 476/ HWC 1477 | OWD 55/ HWC 44 | OWD 52.9/ HWC 51.4 | OWD 25.1/ HWC 24.7 | Serum | Enzyme immunoassay | Korea | Y | N | WHO | NA | Y |
| Lee/ 2013 | IL-18 | OWD 47/ HWC 57 | OWD 66/ HWC 68 | OWD 51.9/ HWC 52.0 | OWD 25.4/ HWC 23.4 | Serum | ELISA | South Korea | N | NA | ADA | 0 | NA |
| Li/ 2020 | IL-17 | LD 40/ HWC 42 | LD 60/ HWC 60 | LD 57.1/ HWC 55.4 | LD 23.85/ HWC 22.28 | Serum | ELISA | China | NA | N | Prevention and Treatment of T2D in China (2017 Edition) | NA | Y |
| Lu/ 2011 | IL-6 | OWD 42/ HWC 41 | OWD 60/ HWC 63 | OWD 55.1/ HWC 48.7 | OWD 27.3/ HWC 24.7 | Plasma | ELISA | Canada | N | N | Canadian Diabetes Association guideline | 10.8 | Y |
| Lukic/ 2014 | IL-6 | OBD 30/ HWC 15 | OBD 50/ HWC 47 | OBD 57.67/ HWC 44.06 | OBD 30.92/ HWC 22.77 | Plasma | ELISA | Serbia | Y | N | WHO | 4.44 | Y |
| Mabrouk/ 2013 | IL-18 | OBD 24/ HWC 30 | OBD 33/ HWC 40 | OBD 32.4/ HWC 30.4 | OBD 53.7/ HWC 22.3 | Serum | ELISA | Egypt | NA | N | NA | NA | NA |
| Malkani/ 2018 | IL-10 | OBD 100/ HWC 100 | NA | OBD 47.9/ HWC 34.3 | OBD 30.11/ HWC 23.52 | Plasma | ELISA | Pakistan | N | N | NA | NA | NA |
| Mishra/ 2011 | IL-6, IL-12 | LD 50/ HWC 50 | NA | LD 46.24/ HWC 43.76 | LD 23.92/ HWC 23.52 | Serum | Cytometric beads array Flex set | India | NA | N | ADA | NA | Y |
| Mohamed/ 2016 | IL-18 | OBD 15/ HWC 10 | NA | OBD 45.0/ HWC 37.8 | OBD 34.0/ HWC 22.2 | Serum | ELISA | Egypt | NA | N | NA | NA | Y |
| Nikbakht/ 2021 | IL-6, IL-8, IL-18 | OBD 12/ HWC 14 | OBD 67/ HWC 57 | OBD 57.7/ HWC 35.2 | OBD 32.59/ HWC 23.36 | Serum | ELISA (IL-6), Human Magnetic Luminex® Assay kit (IL-8, IL-18) | Australia | N | N | WHO | NA | Y |
| Oberbach/ 2006 | IL-6, IL-10 | OBD 20/ HWC 20 | OBD 55/ HWC 45 | NA | OBD 31.3/ HWC 24.2 | Plasma | ELISA | Germany | Y | N | ADA | NA | Y |
| Pedersen/ 2003 | IL-6 | OBD 16/ HWC 20 | NA | OBD 71.5/ HWC 26.5 | OBD 30.25/ HWC 21.95 | Plasma | ELISA | Denmark | Y | Y | WHO | NA | Y |
| Pham/ 2011 | IL-6 | OBD 465/ HWC 41 | OBD 57/ HWC 39 | OBD 56.3/ HWC 47.7 | OBD 30.3/ HWC 23.5 | Serum | Multiplex-bead assay | Germany | NA | NA | NA | 0.1 | Y |
| Qu/ 2013 | IL-6 | OWD 43/ LD 37/ HWC 46 | OWD 42/ LD 30/ HWC 33 | OWD 60.86/ LD 61.65/ HWC 58.54 | OWD 27.78/ LD 22.97/ HWC 22.36 | Plasma | ELISA | China | N | N | WHO | NA | Y |
| Rajkovic/ 2014 | IL-6 | OBD 21/ OWD 18/ LD 25/ HWC 15 | OBD 48/ OWD 56/ LD 52/ HWC 47 | NA | OBD 33.4/ OWD 28.3/ LD 23.8/ HWC 22.7 | Plasma | ELISA | Serbia | Y | N | WHO | OBD 5.84/ OWD 7.3/ LD 6.5 | Y |
| Roohi/ 2014 | IL-17 | OWD 38/ HWC 40 | OWD 50/ HWC 55 | OWD 51.14/ HWC 34.98 | OWD 27.39/ HWC 24.58 | Serum | ELISA | Iran | Y | N | NA | NA | Y |
| Ruge/ 2009 | IL-6 | OWD 10/ HWC 10 | OWD 40/ HWC 40 | OWD 61/ HWC 26 | OWD 27.5/ HWC 22.5 | Plasma | Immunoassay | Sweden | Y | N | NA | NA | Y |
| Schober/ 2007 | IL-6 | OBD 13/ HWC 12 | OBD 100/ HWC 100 | OBD 50/ HWC 54 | OBD 34.7/ HWC 24.0 | Serum | ELISA | Germany | Y | NA | NA | NA | NA |
| Shabangu/ 2019 | IL-10 | OBD 11/ HWC 9 | OBD 18/ HWC 33 | OBD 54.06/ HWC 45.56 | OBD 31.69/ HWC 22.36 | Serum | Luminex multiplex immunoassay | South Africa | N | N | NA | NA | NA |
| Techatanawat/ 2020 | IL-18 | OWD 49/ HWC 25 | OWD 37/ HWC 16 | OWD 61.0/ HWC 54.0 | OWD 25.97/ HWC 23.61 | Serum | ELISA | Thailand | Y | N | NA | NA | Y |
| van Beek/ 2014 | IL-2, IL-4, IL-6, IL-8, IL-10 | OBD 28/ HWC 12 | OBD 0/ HWC 0 | OBD 51/ HWC 50 | OBD 41.7/ HWC 21.7 | Serum | Mesoscale Discovery commercial kit | Netherlands | Y | N | NA | NA | Y |
| Weigert/ 2010 | IL-6 | OBD 30/ HWC 23 | OBD 100/ HWC 100 | OBD 66/ HWC 54 | OBD 30.7/ HWC 23.9 | Serum | ELISA | Germany | Y | NA | NA | NA | Y |
| Woo/ 2012 | IL-6 | OWD 76/ HWC 1224 | OWD 50/ HWC 45 | OWD 56.6/ HWC 49.8 | OWD 25.8/ HWC 23.7 | Serum | ELISA | China | N | N | WHO | 5.3 | NA |
| Zaharieva/ 2018 | IL-6, IL-18 | OBD 76/ HWC 14 | OBD 43/ HWC 7 | OBD 54/ HWC 53 | OBD 33.10/ HWC 21.72 | Serum | Electro-chemiluminescence immunoassay (IL-6), ELISA (IL-18) | Bulgaria | NA | NA | NA | 4 | Y |
| Zhang/ 2014 | IL-2, IL-4, IL-6, IL-10, IL-17 | OWD 32/ HWC 30 | OWD 69/ HWC 67 | OWD 58.47/ HWC 56.90 | OWD 25.18/ HWC 22.12 | Serum | Cytometric beads array | China | NA | N | ADA | 5.06 | Y |
| Zhao/ 2014 | IL-6, IL-17, IL-22 | LD 90/ HWC 30 | LD 52/ HWC 53 | LD 54.36/ HWC 44.43 | LD 24.89/ HWC 22.19 | Plasma | ELISA | China | N | N | NA | 6.14 | Y |
| Zhao/ 2020 | IL-6, IL-17, IL-22 | LD 30/ HWC 20 | LD 60/ HWC 55 | LD 50.37/ HWC 47.10 | LD 24.87/ HWC 21.20 | Plasma | ELISA | China | N | N | NA | 6.34 | Y |
| Zilverschoon/ 2008 | IL-6, IL-18 | OBD 14/ HWC 17 | OBD 57/ HWC 53 | OBD 56.9/ HWC 50.2 | OBD 31.6/ HWC 22.6 | Serum | ELISA | Netherlands | Y | N | NA | 8.3 | Y |

Abbreviation: No., number; T2DM, type 2 diabetes mellitus; HWC, healthy weight control; IL, interleukin; OBD, obese type 2 diabetes mellitus; OWD, overweight type 2 diabetes mellitus; LD, lean type 2 diabetes mellitus; ADA, American Diabetes Association; WHO, World Health Organization; ELISA, enzyme-linked immunosorbent assay; Y, yes; N, no; NA, not available.

Supplemental Table S2. Meta-regression analyses

| Interleukin | Groups | Covariates | No. of Studies | No. With T2D/HWC | Main effect | | | |
| --- | --- | --- | --- | --- | --- | --- | --- | --- |
|  |  |  |  |  | Coefficient (Standard Error) | 95% CI | Z Score | *P*-value |
| IL-2 | Overall | Sex (% Male) | 4 | 142/146 | -0.00616 (0.00549) | -0.01693 to 0.0046 | -1.12236 | 0.2617 |
|  |  | Age | 4 | 142/146 | -0.01234 (0.04981) | -0.10997 to 0.08528 | -0.24779 | 0.8043 |
|  |  | Publication year | 4 | 142/146 | 0.02089 (0.05908) | -0.09489 to 0.13668 | 0.35367 | 0.7236 |
| IL-4 | Overall | Sex (% Male) | 4 | 196/191 | -0.00452 (0.0092) | -0.02256 to 0.01351 | -0.49151 | 0.6231 |
|  |  | Age | 4 | 196/191 | -0.10051 (0.05341) | -0.20519 to 0.00418 | -1.88168 | 0.0599 |
|  |  | Publication year | 4 | 196/191 | -0.35867 (0.07584) | -0.50732 to -0.21003 | -4.72923 | ***<0.0001*** |
|  | *OWD* | Sex (% Male) | 3 | 168/179 | -0.03322 (0.00739) | -0.04771 to -0.01872 | -4.49161 | ***<0.0001*** |
|  |  | Age | 3 | 168/179 | -0.13512 (0.04624) | -0.22576 to -0.04449 | -2.92200 | ***0.0035*** |
|  |  | Publication year | 3 | 168/179 | -0.35915 (0.07798) | -0.51199 to -0.20631 | -4.60564 | ***<0.0001*** |
| IL-6 | Overall | Sex (% Male) | 26 | 1705/3275 | -0.0042 (0.00807) | -0.02002 to 0.01162 | -0.52051 | 0.6027 |
|  |  | Age | 26 | 1725/3280 | -0.02834 (0.02356) | -0.07452 to 0.01785 | -1.20247 | 0.2292 |
|  |  | Publication year | 28 | 1771/3345 | -0.04884 (0.03812) | -0.12356 to 0.02588 | -1.28119 | 0.2001 |
|  | *OBD* | Sex (% Male) | 15 | 790/341 | -0.00642 (0.01058) | -0.02715 to 0.01431 | -0.60687 | 0.5439 |
|  |  | Age | 13 | 739/301 | -0.0512 (0.03614) | -0.12205 to 0.01964 | -1.41668 | 0.1566 |
|  |  | Publication year | 16 | 806/381 | -0.03483 (0.0557) | -0.144 to 0.07434 | -0.62532 | 0.5318 |
|  | *OWD* | Sex (% Male) | 8 | 705/2902 | 0.00923 (0.01503) | -0.02023 to 0.03869 | 0.61402 | 0.5392 |
|  |  | Age | 7 | 687/2887 | 0.03558 (0.08021) | -0.12164 to 0.19279 | 0.44353 | 0.6574 |
|  |  | Publication year | 8 | 705/2902 | 0.15395 (0.0978) | -0.03774 to 0.34565 | 1.57408 | 0.1155 |
|  | *LD* | Sex (% Male) | 6 | 210/190 | 0.00338 (0.00894) | -0.01414 to 0.0209 | 0.37784 | 0.7056 |
|  |  | Age | 6 | 235/225 | -0.01758 (0.0149) | -0.04677 to 0.01162 | -1.17991 | 0.2380 |
|  |  | Publication year | 7 | 260/240 | -0.03872 (0.02725) | -0.09213 to 0.01468 | -1.42130 | 0.1552 |
| IL-8 | Overall | Sex (% Male) | 4 | 130/66 | 0.00349 (0.0076) | -0.01141 to 0.01838 | 0.45892 | 0.6463 |
|  |  | Age | 4 | 130/66 | -0.05854 (0.02859) | -0.11458 to -0.0025 | -2.04730 | ***0.0406*** |
|  |  | Publication year | 4 | 130/66 | 0.12032 (0.06341) | -0.00396 to 0.24459 | 1.89755 | 0.0578 |
|  | *OBD* | Sex (% Male) | 3 | 75/47 | 0.00369 (0.00567) | -0.00744 to 0.01481 | 0.64947 | 0.5160 |
|  |  | Age | 3 | 75/47 | 0.04009 (0.07172) | -0.10047 to 0.18065 | 0.55900 | 0.5762 |
|  |  | Publication year | 3 | 75/47 | 0.05103 (0.07407) | -0.09415 to 0.1962 | 0.68888 | 0.4909 |
| IL-10 | Overall | Sex (% Male) | 6 | 125/133 | -0.02075 (0.01522) | -0.05058 to 0.00907 | -1.36380 | 0.1726 |
|  |  | Age | 5 | 179/168 | -0.16067 (0.07973) | -0.31693 to -0.00441 | -2.01523 | ***0.0439*** |
|  |  | Publication year | 7 | 225/233 | 0.19802 (0.10674) | -0.01118 to 0.40723 | 1.85524 | 0.0636 |
|  | *OBD* | Sex (% Male) | 4 | 85/86 | -0.04589 (0.00722) | -0.06004 to -0.03174 | -6.35720 | ***<0.0001*** |
|  |  | Age | 3 | 139/121 | -0.73713 (0.20514) | -1.1392 to -0.33507 | -3.59337 | ***0.0003*** |
|  |  | Publication year | 5 | 185/186 | 0.31745 (0.11646) | 0.08918 to 0.54572 | 2.72571 | ***0.0064*** |
| IL-12 | Overall | Sex (% Male) | 3 | 155/168 | -0.09335 (0.03369) | -0.15939 to -0.02731 | -2.77057 | ***0.0056*** |
|  |  | Age | 4 | 205/218 | -0.09596 (0.07342) | -0.23986 to 0.04794 | -1.30705 | 0.1912 |
|  |  | Publication year | 4 | 205/218 | -0.598 (0.24303) | -1.07433 to -0.12167 | -2.46060 | ***0.0139*** |
| IL-17 | Overall | Sex (% Male) | 6 | 244/184 | 0.22054 (0.06414) | 0.09482 to 0.34625 | 3.43826 | ***0.0006*** |
|  |  | Age | 6 | 244/184 | 0.76288 (0.15803) | 0.45314 to 1.07262 | 4.82729 | ***<0.0001*** |
|  |  | Publication year | 6 | 244/184 | 0.43621 (0.36941) | -0.28782 to 1.16024 | 1.18083 | 0.2377 |
|  | *LD* | Sex (% Male) | 3 | 160/92 | 0.39767 (0.29488) | -0.18028 to 0.97563 | 1.34859 | 0.1775 |
|  |  | Age | 3 | 160/92 | 0.63554 (0.36836) | -0.08643 to 1.35751 | 1.72534 | 0.0845 |
|  |  | Publication year | 3 | 160/92 | 0.53023 (0.39317) | -0.24038 to 1.30084 | 1.34859 | 0.1775 |
| IL-18 | Overall | Sex (% Male) | 8 | 318/252 | -0.01093 (0.02752) | -0.06487 to 0.04301 | -0.39721 | 0.6912 |
|  |  | Age | 9 | 333/262 | -0.14295 (0.03887) | -0.21914 to -0.06677 | -3.67783 | ***0.0002*** |
|  |  | Publication year | 9 | 333/262 | -0.17848 (0.09147) | -0.35775 to 0.0008 | -1.95124 | 0.0510 |
|  | *OBD* | Sex (% Male) | 5 | 147/100 | -0.00592 (0.03583) | -0.07615 to 0.0643 | -0.16531 | 0.8687 |
|  |  | Age | 6 | 162/110 | -0.12909 (0.04963) | -0.22636 to -0.03183 | -2.60138 | ***0.0093*** |
|  |  | Publication year | 6 | 162/110 | -0.24342 (0.08864) | -0.41714 to -0.0697 | -2.74629 | ***0.0060*** |
|  | *OWD* | Sex (% Male) | 3 | 171/152 | -0.01465 (0.02467) | -0.06301 to 0.03371 | -0.59384 | 0.5526 |
|  |  | Age | 3 | 171/152 | -0.11999 (0.04599) | -0.21012 to -0.02986 | -2.60925 | ***0.0091*** |
|  |  | Publication year | 3 | 171/152 | -0.15448 (0.05287) | -0.2581 to -0.05086 | -2.92208 | ***0.0035*** |
| IL-22 | Overall | Sex (% Male) | 3 | 395/2806 | 0.09835 (0.01318) | 0.07251 to 0.12419 | 7.46026 | ***<0.0001*** |
|  |  | Age | 3 | 395/2806 | -0.216 (0.02887) | -0.27259 to -0.15941 | -7.48086 | ***<0.0001*** |
|  |  | Publication year | 3 | 395/2806 | -0.04053 (0.16089) | -0.35586 to 0.27481 | -0.25190 | 0.8011 |
| IL-33 | Overall | Publication year | 3 | 118/125 | 0.0944 (0.03759) | 0.02073 to 0.16807 | 2.51134 | ***0.0120*** |

Abbreviation: No., number; CI, confidence interval; T2D, type 2 diabetes; HWC, healthy weight control; IL, interleukin; OBD, obese type 2 diabetes; OWD, overweight type 2 diabetes; LD, lean type 2 diabetes; Z (test of null hypothesis); P (statistical significance)

Supplemental Table S3. PRISMA 2020 Checklist

| **Section and Topic** | **Item #** | **Checklist item** | **Location where item is reported** |
| --- | --- | --- | --- |
| **TITLE** | | |  |
| Title | 1 | Identify the report as a systematic review. | Page (P)1, Line (L)1-2 |
| **ABSTRACT** | | |  |
| Abstract | 2 | See the PRISMA 2020 for Abstracts checklist. | P2-3, L20-45 |
| **INTRODUCTION** | | |  |
| Rationale | 3 | Describe the rationale for the review in the context of existing knowledge. | P3-5, L59-99 |
| Objectives | 4 | Provide an explicit statement of the objective(s) or question(s) the review addresses. | P5, L101-105 |
| **METHODS** | | |  |
| Eligibility criteria | 5 | Specify the inclusion and exclusion criteria for the review and how studies were grouped for the syntheses. | P6, L118-129 |
| Information sources | 6 | Specify all databases, registers, websites, organisations, reference lists and other sources searched or consulted to identify studies. Specify the date when each source was last searched or consulted. | P6, L113-114 |
| Search strategy | 7 | Present the full search strategies for all databases, registers and websites, including any filters and limits used. | P6, L115-116 |
| Selection process | 8 | Specify the methods used to decide whether a study met the inclusion criteria of the review, including how many reviewers screened each record and each report retrieved, whether they worked independently, and if applicable, details of automation tools used in the process. | P6, L113-130 |
| Data collection process | 9 | Specify the methods used to collect data from reports, including how many reviewers collected data from each report, whether they worked independently, any processes for obtaining or confirming data from study investigators, and if applicable, details of automation tools used in the process. | P7, L132-143 |
| Data items | 10a | List and define all outcomes for which data were sought. Specify whether all results that were compatible with each outcome domain in each study were sought (e.g. for all measures, time points, analyses), and if not, the methods used to decide which results to collect. | P7, L132-143 |
|  | 10b | List and define all other variables for which data were sought (e.g. participant and intervention characteristics, funding sources). Describe any assumptions made about any missing or unclear information. | P7, L134-136 |
| Study risk of bias assessment | 11 | Specify the methods used to assess risk of bias in the included studies, including details of the tool(s) used, how many reviewers assessed each study and whether they worked independently, and if applicable, details of automation tools used in the process. | P7, L137-139 |
| Effect measures | 12 | Specify for each outcome the effect measure(s) (e.g. risk ratio, mean difference) used in the synthesis or presentation of results. | P7, L148-151 |
| Synthesis methods | 13a | Describe the processes used to decide which studies were eligible for each synthesis (e.g. tabulating the study intervention characteristics and comparing against the planned groups for each synthesis (item #5)). | P6, L120-123 |
|  | 13b | Describe any methods required to prepare the data for presentation or synthesis, such as handling of missing summary statistics, or data conversions. | P7, L148-149 |
|  | 13c | Describe any methods used to tabulate or visually display results of individual studies and syntheses. | P7, L149-152 |
|  | 13d | Describe any methods used to synthesize results and provide a rationale for the choice(s). If meta-analysis was performed, describe the model(s), method(s) to identify the presence and extent of statistical heterogeneity, and software package(s) used. | P7, L145-146, L152-154; P8, L158-161 |
|  | 13e | Describe any methods used to explore possible causes of heterogeneity among study results (e.g. subgroup analysis, meta-regression). | P7, L146-148; P8, L161-164 |
|  | 13f | Describe any sensitivity analyses conducted to assess robustness of the synthesized results. | P7-8, L154-156 |
| Reporting bias assessment | 14 | Describe any methods used to assess risk of bias due to missing results in a synthesis (arising from reporting biases). | P8, L164-169 |
| Certainty assessment | 15 | Describe any methods used to assess certainty (or confidence) in the body of evidence for an outcome. | P7-8, L154-169 |
| **RESULTS** | | |  |
| Study selection | 16a | Describe the results of the search and selection process, from the number of records identified in the search to the number of studies included in the review, ideally using a flow diagram. | P8-9, L176-180 & Fig. 1 |
|  | 16b | Cite studies that might appear to meet the inclusion criteria, but which were excluded, and explain why they were excluded. | Fig. 1 |
| Study characteristics | 17 | Cite each included study and present its characteristics. | P6, L129 & Table S1 |
| Risk of bias in studies | 18 | Present assessments of risk of bias for each included study. | Table 1 |
| Results of individual studies | 19 | For all outcomes, present, for each study: (a) summary statistics for each group (where appropriate) and (b) an effect estimate and its precision (e.g. confidence/credible interval), ideally using structured tables or plots. | Figs 2 & 3 |
| Results of syntheses | 20a | For each synthesis, briefly summarise the characteristics and risk of bias among contributing studies. | Table 1 |
|  | 20b | Present results of all statistical syntheses conducted. If meta-analysis was done, present for each the summary estimate and its precision (e.g. confidence/credible interval) and measures of statistical heterogeneity. If comparing groups, describe the direction of the effect. | Table 1 |
|  | 20c | Present results of all investigations of possible causes of heterogeneity among study results. | Table 1, Table S2 |
|  | 20d | Present results of all sensitivity analyses conducted to assess the robustness of the synthesized results. | Table 1 |
| Reporting biases | 21 | Present assessments of risk of bias due to missing results (arising from reporting biases) for each synthesis assessed. | Table 1 |
| Certainty of evidence | 22 | Present assessments of certainty (or confidence) in the body of evidence for each outcome assessed. | Table 1 |
| **DISCUSSION** | | |  |
| Discussion | 23a | Provide a general interpretation of the results in the context of other evidence. | P12-16, L251-342 |
|  | 23b | Discuss any limitations of the evidence included in the review. | P16, L345-350 |
|  | 23c | Discuss any limitations of the review processes used. | P16, L354-356 |
|  | 23d | Discuss implications of the results for practice, policy, and future research. | P12-16, L266-342 |
| **OTHER INFORMATION** | | |  |
| Registration and protocol | 24a | Provide registration information for the review, including register name and registration number, or state that the review was not registered. | P5, L109 |
|  | 24b | Indicate where the review protocol can be accessed, or state that a protocol was not prepared. | P5, L109 |
|  | 24c | Describe and explain any amendments to information provided at registration or in the protocol. | Not applicable |
| Support | 25 | Describe sources of financial or non-financial support for the review, and the role of the funders or sponsors in the review. | P17, L378-381 |
| Competing interests | 26 | Declare any competing interests of review authors. | P17, L382 |
| Availability of data, code and other materials | 27 | Report which of the following are publicly available and where they can be found: template data collection forms; data extracted from included studies; data used for all analyses; analytic code; any other materials used in the review. | Table S1 |
